# Changing transmission dynamics of COVID-19 in China: a nationwide population-based piecewise mathematical modelling study

**DOI:** 10.1101/2020.03.27.20045757

**Authors:** Jiawen Hou, Jie Hong, Boyun Ji, Bowen Dong, Yue Chen, Michael P. Ward, Wei Tu, Zhen Jin, Jian Hu, Qing Su, Wenge Wang, Zheng Zhao, Shuang Xiao, Jiaqi Huang, Wei Lin, Zhijie Zhang

**Affiliations:** Institute of Science and Technology for Brain-Inspired Intelligence, Fudan University, Shanghai 200433, China; Centre for Computational Systems Biology and Research Institute of Intelligent Complex Systems, Fudan University, Shanghai 200433, China; Department of Epidemiology and Health Statistics, School of Public Health, Fudan University, Shanghai 200032, China; Key Laboratory of Public Health Safety, Ministry of Education, Shanghai 200032, China; School of Mathematical Sciences and Shanghai Center for Mathematical Sciences, Fudan University, Shanghai 200433, China; Department of Epidemiology and Community Medicine, Faculty of Medicine, University of Ottawa, 451 Smyth Rd, Ottawa, Ontario, Canada; Faculty of Veterinary Science, The University of Sydney NSW, Sydney, Australia; Department of Geology and Geography, Georgia Southern University, Statesboro, GA 30460, USA; Complex Systems Research Center, Shanxi University, Taiyuan, Shan’xi 030006; Shanghai Key Laboratory for Contemporary Applied Mathematics, LNMS (Fudan University), and LCNBI (Fudan University), Shanghai 200433, China; State Key Laboratory of Medical Neurobiology, and MOE Frontiers Center for Brain Science, Institutes of Brain Science, Fudan University, Shanghai 200032, China

**Keywords:** COVID-19, Multi-stage SEIR model, Source tracing algorithm, Controlled reproduction number, Heterogeneity, Epidemic peak

## Abstract

**Background:** The first case of COVID-19 atypical pneumonia was reported in Wuhan, China on December 1, 2019. Since then, at least 33 other countries have been affected and there is a possibility of a global outbreak. A tremendous amount of effort has been made to understand its transmission dynamics; however, the temporal and spatial transmission heterogeneity and changing epidemiology have been mostly ignored. The epidemic mechanism of COVID-19 remains largely unclear.

**Methods:** Epidemiological data on COVID-19 in China and daily population movement data from Wuhan to other cities were obtained and analyzed. To describe the transmission dynamics of COVID-19 at different spatio-temporal scales, we used a three-stage continuous-time Susceptible-Exposed-Infectious-Recovered (SEIR) meta-population model based on the characteristics and transmission dynamics of each stage: 1) local epidemic from December 1, 2019 to January 9, 2020; 2) long-distance spread due to the Spring Festival travel rush from January 10 to 22, 2020; and 3) intra-provincial transmission from January 23, 2020 when travel restrictions were imposed. Together with the basic reproduction number (*R*_0_) for mathematical modelling, we also considered the variation in infectivity and introduced the controlled reproduction number (*R*_*c*_) by assuming that exposed individuals to be infectious; we then simulated the future spread of COVID across Wuhan and all the provinces in mainland China. In addition, we built a novel source tracing algorithm to infer the initial exposed number of individuals in Wuhan on January 10, 2020, to estimate the number of infections early during this epidemic.

**Findings:** The spatial patterns of disease spread were heterogeneous. The estimated controlled reproduction number (*R*_*c*_) in the neighboring provinces of Hubei province were relatively large, and the nationwide reproduction number ‐ except for Hubei ‐ ranged from 0.98 to 2.74 with an average of 1.79 (95% CI 1.77‐1.80). Infectivity was significantly greater for exposed than infectious individuals, and exposed individuals were predicted to have become the major source of infection after January 23. For the epidemic process, most provinces reached their epidemic peak before February 10, 2020. It is expected that the maximum number of infections will be approached by the end of March. The final infectious size is estimated to be about 58,000 for Wuhan, 20,800 for the rest of Hubei province, and 17,000 for the other provinces in mainland China. Moreover, the estimated number of the exposed individuals is much greater than the officially reported number of infectious individuals in Wuhan on January 10, 2020.

**Interpretation:** The transmission dynamics of COVID-19 have been changing over time and were heterogeneous across regions. There was a substantial underestimation of the number of exposed individuals in Wuhan early in the epidemic, and the Spring Festival travel rush played an important role in enhancing and accelerating the spread of COVID-19. However, China’s unprecedented large-scale travel restrictions quickly reduced *R*_*c*_. The next challenge for the control of COVID-19 will be the second great population movement brought by removing these travel restrictions.

## Introduction

On December 1, 2019, a case of pneumonia of unknown etiology was reported in Wuhan, China, and a novel strain of coronavirus (SARS-CoV-2) associated with the pneumonia case was subsequently isolated on January 7, 2020^1,2^. As of February 24, 2020, about 78,000 cases had been confirmed (including clinically diagnosed cases) in China and 33 other countries, of which about 65,000 were from Hubei province. There had been 2,666 deaths in China, of whom most were elderly people or people having underlying health conditions.

While sustained human-to-human transmission of SARS-CoV-2 is clear, there remain many questions about specific transmission characteristics of the virus^3^. Because disease control efforts change dramatically over time and vary across geographic regions, the epidemic of COVID-19 shows temporal and spatial transmission patterns. The few studies to date on transmission modelling and forecasting of COVID-19 have estimated the basic reproduction number (*R*_0_) based on limited data in the early stage of the COVID-19 epidemic^4-8^, commonly with an assumption of no changes in the dynamics of transmission, surveillance and control of the epidemic. Transmission and disease control change over time and the epidemic mechanism of COVID-19 still remains largely unclear.

In this study, we developed a spatial-temporal transmission model for the spread of COVID-19 in China. We characterized the transmission dynamics in three epidemic stages defined by population movement (Lunar New Year (Spring Festival)), implementation of control measures and location. Instead of applying the commonly-used *R*_0_ metric to describe infectivity, we estimated the controlled reproduction number, *R*_*c*_, and forecasted the future trend of this epidemic for different provinces of mainland China. We also retrospectively estimated the number of individuals infected in the early stage of the epidemic when under-reporting was common.

## Materials and Methods

### Data sources

#### Epidemiological data

The daily cumulative numbers of confirmed and suspected COVID-19 cases for the period from December 1, 2019 to February 8, 2020 were obtained from the National, Hubei Provincial and Wuhan Municipal Health and Family Planning Commissions. Due to a concern of under-reporting before January 20, 2020, data in this period were imputed and double-checked manually based on a range of sources, including local government reports, officially published literature and local hospital reports^9^.

The definitions of confirmed and suspected cases for COVID-19 have been described elsewhere in detail^8^. We only used data from confirmed cases in the modelling and included all provinces in mainland China except for Tibet and Xinjiang where only a few confirmed cases were reported.

#### Population movement data

As the capital city of Hubei Province, Wuhan has more than 11 million residents and is connected to other major cities in China via frequent buses, high-speed trains, and commercial flights. It is the busiest traffic hub of central China. The daily population movement from and to Wuhan increased dramatically before the Lunar New Year (starting around January 10, 2020), which accelerated the spread of COVID-19^10,11^. The daily population movement data into and out of Wuhan by types of transportation (i.e., long-distance bus, train and air) for the period January 10‐23, 2020 were retrieved from an app-derived real-time location database provided by Baidu Huiyan Company, from which we constructed a travel volume matrix among different provinces.

#### A three-stage piecewise mathematical model

We established a continuous-time but piecewise Susceptible-Exposed-Infectious-Recovered (SEIR) meta-population model to study the transmission dynamics of COVID-19. We considered transmission dynamics separately for each province. We modelled Wuhan city separately because of its heavy disease burden; other cities of Hubei province were modelled together. We divided the epidemic of COVID-19 into three developmental stages and the first two stages were mainly focused on Wuhan, whereas in the last stage we considered epidemics in all provinces.

Stage 1 commenced on December 1, 2019 when the first COVID-19 case was detected, and ended on January 8, 2020, prior to the mass Lunar New Year population movement. During this period, the disease mainly spread within Wuhan, and inter-provincial movement of infected people was ignored. The transmission dynamics are described by the following differential equations:

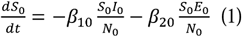

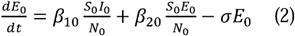

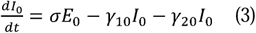

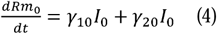

where *N*_0_ denotes the population size of Wuhan, *S*_0_, *E*_0_, *I*_0_ and *Rm*_0_ denotes the number of susceptible, exposed (incubation), symptomatic and removed individuals, respectively. Although “*I*” represents infectious individuals in traditional SEIR models, we assumed all exposed individuals were also infectious and used *I*_0_ to represent the symptomatic phase. As it is not clear how prevalent is asymptomatic infection, we assumed for model simplicity that all exposed individuals move to the symptomatic phase. The infectiousness levels of exposed and symptomatic individuals are represented by the infection forces *β*_10_ and *β*_20_, respectively, and *σ* represents the rate of developing symptoms among exposed people. Previous studies on nonspecific pneumonia have assumed *σ* to be 1/7, which implies an average incubation period of one week^12^. The removed state *R*_0_ includes both recovered and decreased patients, with *γ*_10_and *γ*_20_ representing the recovery and mortality rates, respectively.

Stage 2 of the epidemic from January 10 to 22, 2020 covered the period of mass migration of people returning to their hometown for the Lunar New Year. Few cases were detected outside Wuhan during this period, and therefore we still focused on Wuhan but added a migration term to reflect outbound human movement:

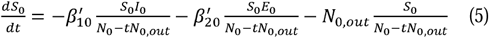

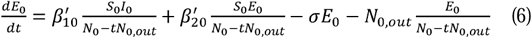

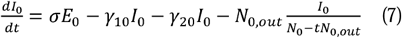

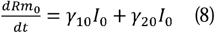

where *N*_0,*out*_ denotes the emigration rate, measured as the number of emigration events per person per day from Wuhan. The emigration of recovered individuals was ignored as such movement has negligible impact on the model. Due to potentially increased social interactions during this period, the infection forces 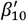 and 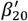 differ from those in stage 1.

Stage 3 commenced from January 23, 2020 when a lockdown was in effect in Wuhan. Due to the strictly enforced travel ban, inter-provincial human movement was significantly reduced. Therefore, we assumed that the transmission dynamics in each province evolved independently. Here, Hubei province (excluding Wuhan) was regarded as a whole region. Considering that the tightened control policies reduced the contact frequencies of residents within each province, we added an attenuation effect *λ(α*_i_, *t)* to the infection risk from exposed individuals in their incubation period, where *α*_i_ is the attenuation coefficient specific to the *i*-th province. The model in this stage is desribed by:

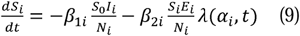

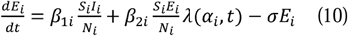

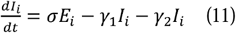

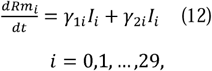

where the subsript *i* indicates the *i*-th province, with *i* = 0 designated for Wuhan and *i* = 1 for the remaining cities of Hubei as a whole. Each province has its own infection forces. Recovery rate and mortality rate also differ by province, which is reasonable due to the heterogeneity in medical resources. The only parameter shared by all provinces is *σ*, the rate of disease development which depends more on the pathogen than on the population. Considering the fact that medical resources in Wuhan were exhausted first but improved later on with aid from other parts of the country, we add an enhancement term *µ*(*α*_20_, *t*) to the case recovery rate:

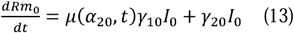

### Estimation and prediction

The key parameters in the above models were estimated using MATLAB R2018b through the least square technique. The 95% confidence interval (95% CI) of the parameter was estimated by independently adding random terms to the initial parameter value during the fitting process. The models for different provinces were calibrated separately. To assess the goodness-of-fit of the models, we calculated correlation coefficients between simulated and observed data, i.e., between model-predicted cumulative number of infections and the reported cumulative numbers of confirmed cases by each day. The fitted model was then used to predict the outbreak size and the timing of outbreak peak in each province, assuming the control policies were time-invariant until the outbreak ends. The epidemic peak for each province was defined as the day on which the estimated number of new cases reached a maximum.

For Stage 1 without variation in transmission and recovery rates, we use the conventional method to calculate the basic reproduction number (*R*_0_) in Wuhan for our model approximately at the disease free equilibrium^13^:

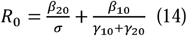

For Stage 2 in Wuhan, we modify the above calculation considering the population movement:

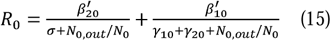

For Stage 3, since some effects dependent on time evolution are taken into account not only in equation (10) for all provinces but also in equation (13) for Wuhan, we obtained the reproduction numbers as functions of time *t* as follows:

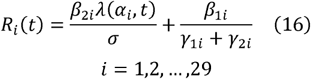

and

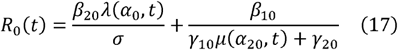

The median *R*_*i*_*(t)* was used to evaluate the average of the effective infection rate for Wuhan and other provinces, respectively, which we refer to as the median controlled reproduction number *R*_*c*_ ^14^.

### Estimating the exposed population size in Wuhan in the early epidemic phase

To retrospectively identify the number of exposed individuals in Wuhan on January 10, we developed a simplified SE model to describe the transmission among the migrating population from Wuhan to the other provinces as follows:

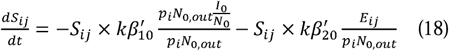

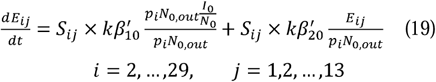

where *S*_*ij*_ and *E*_*ij*_ are the numbers of susceptible and exposed individuals, respectively, who traveled from Wuhan to the *i*-th province on the *j*-th day during Stage 2. In addition, *p*_*i*_ represents the proportion of individuals who traveled to the *i*-th province, and *k* is a correction factor for infectivity given that the contacts in transportation vehicles are likely to be much closer than in the general society. In this model, we used parameters estimated in Stage 2 for Wuhan and the numbers of exposed individuals in the other provinces at the beginning of Stage 3 to infer the number of exposed individuals on January 10 in Wuhan, i.e. back-estimating in time. The numbers of exposed individuals in the other provinces at the beginning of Stage 3 were calculated from the epidemiological data.

### Ethics Approval

The National Health Commission of China determined that the collection of data from human cases of COVID-19 was part of a continuing public health surveillance of a notifiable infectious disease and was thus exempt from institutional review board approval.

### Results

The stage-specific and overall correlations between model predicted and observed cumulative case numbers were >0.9 for all the provinces with the exception of Wuhan in Stage 1 (Table 1 and in Supplementary Table 1), which was 0.785. The less satisfactory correlation for the first stage in Wuhan city is likely caused by under-reporting during the early epidemic phase.

**Table 1.**
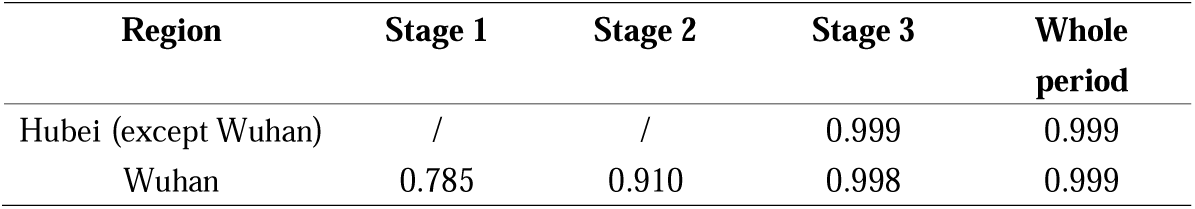
The estimation accuracies of the model for Wuhan and the Hubei province (except Wuhan), respectively.

| Region | Stage 1 | Stage 2 | Stage 3 | Whole period |
| --- | --- | --- | --- | --- |
| Hubei (except Wuhan) | / | / | 0.999 | 0.999 |
| Wuhan | 0.785 | 0.910 | 0.998 | 0.999 |

Estimates of the key parameters in each epidemic stage are shown in Table 2 for Wuhan and the rest of Hubei province. The results demonstrate that the estimated *R*_0_ dropped drastically in Stage 2 for Wuhan and the estimated median controlled reproduction number (*R*_*c*_) was significantly higher in Wuhan in Stage 3 compared to th rest of Hubei. In Stage 3, the infectivity of exposed individuals (*β*_20_) was significantly higher than that of the infectious individuals for both Wuhan and the rest of Hubei, suggesting that the exposed individuals had become the major source of infection after January 23, 2020.

**Table 2.** Key parameters estimated for the city of Wuhan and the Hubei province (except Wuhan), respectively.

| Region | Stage | $\beta_1$ | $\beta_2$ | $\gamma_1$ | $\gamma_2$ | $\alpha_1$ | $\alpha_2$ | $R_0(R_c)$<br>(95% CI) |
| --- | --- | --- | --- | --- | --- | --- | --- | --- |
| Wuhan | 1 | 1.39E-01 | 1.54E-01 | 3.64E-02 | 2.94E-05 | / | / | 4.90<br>(3.32, 6.20) |
|  | 2 | 1.59E-01 | 1.81E-01 | 3.64E-02 | 2.94E-05 | / | / | 3.00<br>(2.21, 4.37) |
|  | 3 | 5.00E-02 | 6.59E-01 | 4.38E-03 | 1.01E-02 | 1.36E-01 | 1.10E-01 | 4.75<br>(4.49, 4.98) |
| Hubei<br>except<br>Wuhan | 3 | 2.22E-14 | 6.76E-01 | 1.83E-06 | 2.64E-03 | 1.70E-01 |  | 2.07 ( 1.97,2.09 ) |

We mapped the spatial distribution of province-specific *R*_*c*_ estimates (Figure 1) and other key parameter estimates (Supplementary Figure 1). Hubei province and its neighboring provinces, together with Hebei, Gansu, Jilin and Heilongjiang provinces, had relatively higher values of *R*_*c*_. The nationwide controlled reproduction number (excluding Hong Kong, Macau, Taiwan, Xinjiang, Tibet, and Hubei) was 1.79 (95% CI 1.77‐1.80). The detailed dereasing process for *R*_*i*_*(t)* in Wuhan, other cities of Hubei and other provinces is illustrated in Supplementary Figure 2; for most provinces *R*_*i*_*(t)* reached the threshold of 1 before February 16, 2020 (Supplementary Table 4), at which point the epidemic was predicted to be fading out.

**Figure 1:**
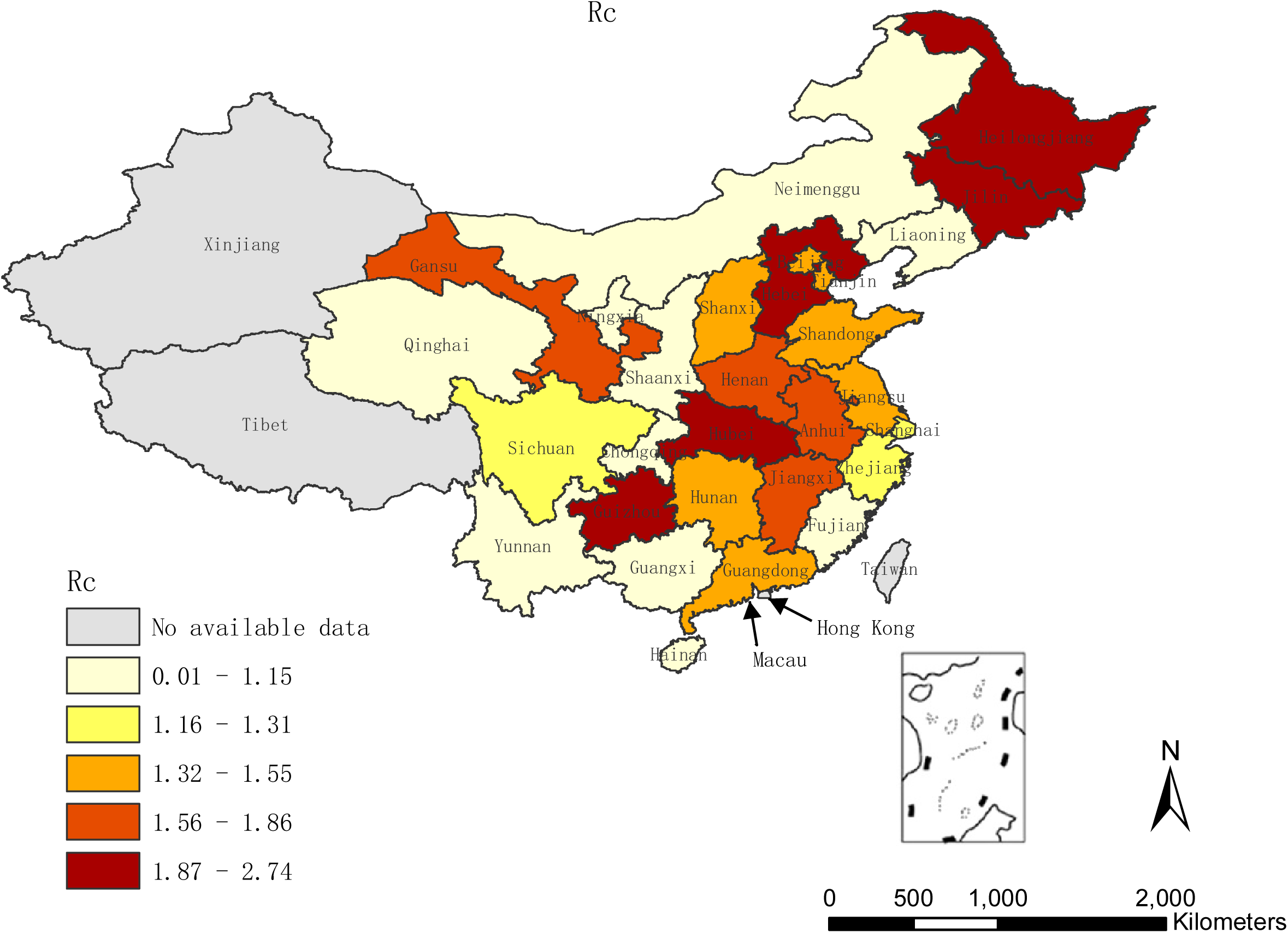
The median controlled reproduction number of COVID-19 for all provinces in mainland China. Tibet and Xinjiang are excluded because of a lack of available data.

Using the model-estimated parameters, we simulated the epidemic from February 8, 2020, to predict the cumulative number of cases occurring by March 31, 2020, before the point in time most provinces were expected to reach their epidemic process endpoints. The models predicted a final size of infected COVID-19 cases of about 58,000 in Wuhan, 20,500 in the rest of Hubei, and 17,000 in all other provinces of mainland China (Figure 2, Supplementary Figure 3 and Supplementary Table 3). Most provinces have reached their epidemic peak before February 10, 2020(Figure 3). The number of new COVID-19 cases is expected to continually decrease if no other special events occur.

**Figure 2:**
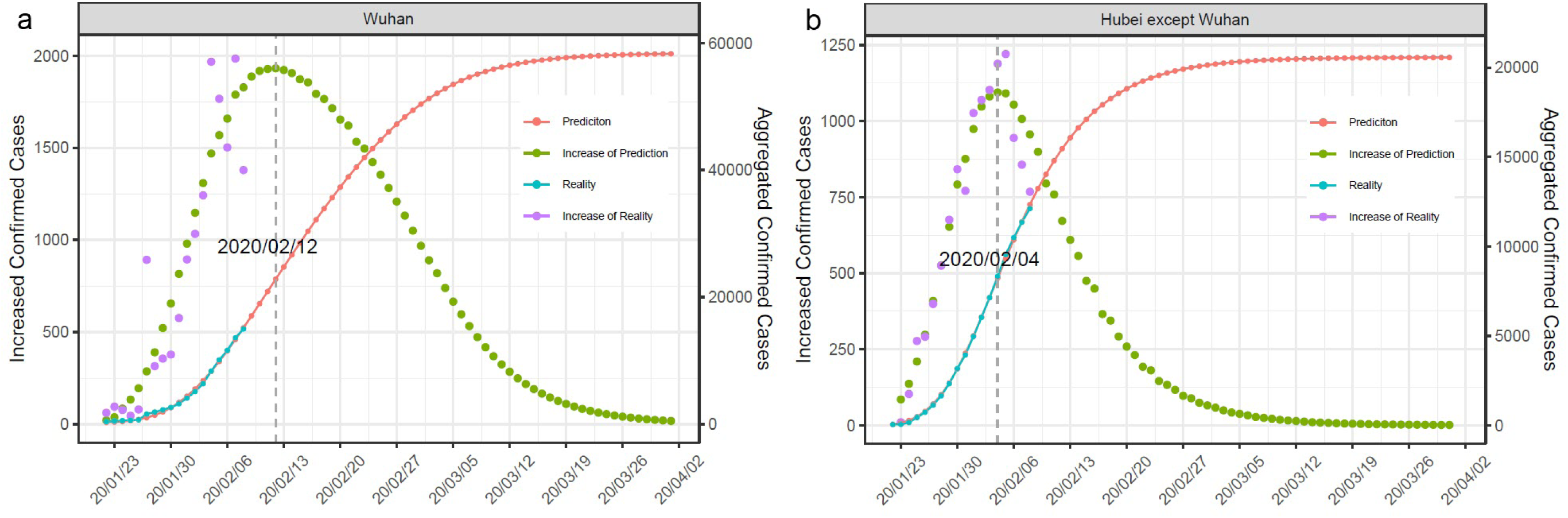
The predicted infectious sizes for COVID-19 in Wuhan (a) and other cities of Hubei (b). Red: the predicted total infectious cases; Green: the predicted new infectious cases; Blue: the reported total infectious cases; Purple: the reported new infectious numbers.

**Figure 3:**
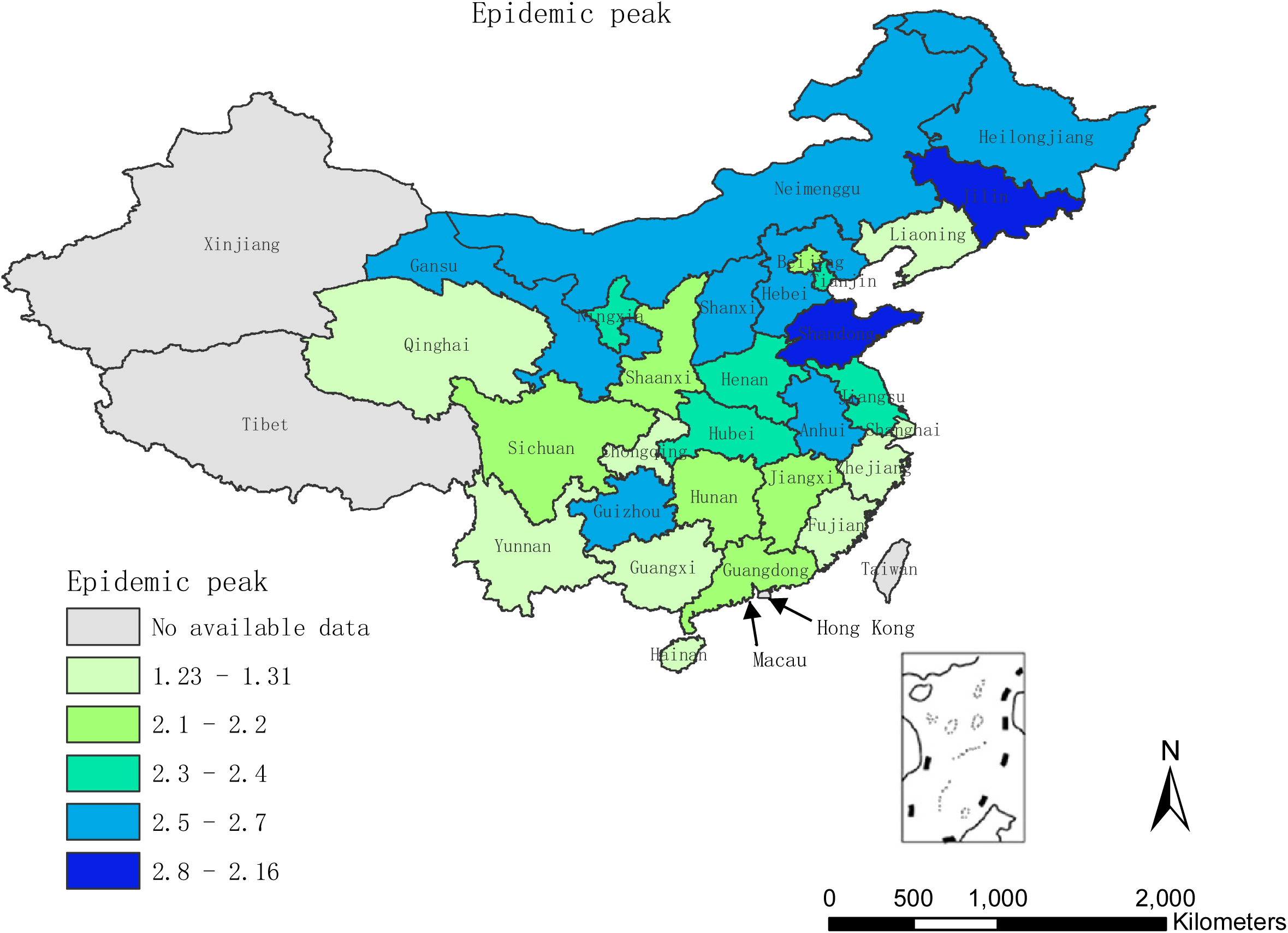
The COVID-19 epidemic peaks for all provinces in mainland China, except for Tibet and Xinjiang.

To address the concern of under-reporting in Wuhan in Stage 1, we proposed a source tracing strategy implemented through transportation in Stage 2. With the estimated initial exposed individuals on January 23, 2020, the model is able to attain the epidemic situation in Stage 3 (Supplementary Table 3). The estimated number of exposed individuals on January 10, 2020 in Wuhan is a function of the correction factor *k* and the number of infected individuals on January 10, 2020, *I*_0_. As is shown in Table 3, the number of initial exposed individuals decreases significantly as *k* increases, and decreases slightly at a linear trend as *I*_0_ increases. The estimated number of exposed individuals is much greater than the reported infectious individuals on January 17, 2020 (7 days or an incubation period after January 10, 2020) in Wuhan.

**Table 3.** Estimated exposed numbers in Wuhan on January 10, 2020 with different pairs of values for two variables *k* and *I*_0_.

| $I_0 \backslash k$ | **40** | **45** | **50** | **55** | **60** |
| --- | --- | --- | --- | --- | --- |
| **0.5** | 2163 | 2159 | 2155 | 2152 | 2148 |
| **0.75** | 2032 | 2028 | 2025 | 2021 | 2017 |
| **1** | 1911 | 1907 | 1903 | 1900 | 1896 |
| **1.25** | 1798 | 1795 | 1791 | 1787 | 1784 |
| **1.5** | 1694 | 1690 | 1686 | 1683 | 1679 |
| **1.75** | 1596 | 1593 | 1589 | 1585 | 1582 |
| **2** | 1506 | 1502 | 1498 | 1495 | 1491 |
| **2.25** | 1421 | 1417 | 1414 | 1410 | 1406 |
| **2.5** | 1342 | 1338 | 1334 | 1331 | 1327 |
| **2.75** | 1268 | 1264 | 1260 | 1256 | 1253 |
| **3** | 1198 | 1195 | 1191 | 1187 | 1183 |

## Discussion

The spatial pattern of the transmission dynamics of COVID-19 shows a strong heterogeneity. The provinces close to the Hubei province-such as Hunan, Jiangxi and Henan-generally had relatively larger values of *R*_*c*_ (Figure. 1). The population movement from Wuhan to these provinces were relatively higher compared to other provinces, therefore so too were more infectious individuals. Heilongjiang and Jilin in northeast China, and some provinces (e.g., Gansu and Guizhou) had relatively large values of *R*_*c*_, for which there is currently no clear explanation. Further investigation of local disease spread conditions are warranted.

The dynamics of the epidemic was also time-varying, and the commonly-used basic reproduction number *R*_0_ does not take this variation of infectivity into account. We modelled R(t), which gradually decreased over time in Stage 3. The R(t) for Wuhan was about 8.1 on January 23, 2020, and crossed the threshold on <1 on February 17, 2020 as a result of the intervention measures applied. The infectivity of exposed and symptomatic individuals varied across the different epidemic stages in Wuhan. In epidemic stages 1 and 2, before the lockdown was in effect in Wuhan, the transmission rates for exposed and symptomatic individuals (*β*_10_ and *β*_20_, respectively) were about equal. However, in stage 3 the infectivity of exposed individuals was significantly greater than that of the infectious individuals, which was the case for most of the other provinces. When symptomatic individuals were isolated and treated in time, exposed individuals then became the major source of infection after January 23, 2020.

We found that the estimated *R*_*c*_ value for COVID-19 in Wuhan was significantly higher than those reported for SARS and MERS. Since the estimated values are mostly between 2 and 3 for SARS^15^ and are uniformly lower for MERS^16^, COVID-19 can be much more infectious in some situations, confirmed by the current and previous studies^17^. Our estimated *R*_0_ in different epidemic stages also reflect the dynamics of surveillance effort and reporting.

The epidemic peak is defined as the day on which the number of new cases reaches its maximum. Most provinces reached their epidemic peaks between February 1 and 16, 2020. The nationwide epidemic peak, excluding Hebei province, occurred on February 3, but there are some uncertainties concerning the potential second great population movement brought by the resumption of work and education 18. We also calculated the maximum number of infectious individuals, which is another key indicator for epidemic control. Our simulation results revealed that most provinces would approach the peak of total confirmed cases before April, assuming that the epidemic in each province evolves independently without a large inter-provincial population movement. Since Spring Festival holidays are largely extended, a huge returning flow of population can invalidate our assumption and therefore model predictions. As a result, a new epidemic wave could happen, which requires close attention from all stakeholders.

The number of cases in Wuhan in epidemic stages 1 and 2 might be substantially underreported due to various reasons. To address this problem, we designed a method to estimate the epidemic scale in the first two stages. Our numerical results demonstrate a large degree of underreporting with a similar conclusion to a previous study^6^ and these exposed individuals likely became a major source of intra- or inter-provincial infection. Underreporting of exposed individuals could lead to an underestimation of infectious individuals in stages 1 and 2. Considering the rapidly increasing number of infectious individuals in Wuhan after January 23, 2020, our estimation of the initial exposed number based on the spatio-temporal piecewise structure is necessary and helpful for understanding the true early situations.

There are several limitations for this study. Firstly, individuals with no or only mild symptoms might not seek treatment, and therefore, not be included in the dataset, especially for epidemic stages 1 and 2. This reporting bias could result in underestimation for the number of cases as well as *R*_0_. Secondly, we did not include epidemic data from Hong Kong, Macau, Taiwan, and foreign countries. Spatial and temporal heterogeneity would have been larger if these data were included. Finally, we used data from laboratory diagnosed cases, and therefore, the number of predicted cases after February 12 would be lower than the officially reported number of both clinically and laboratory diagnosed cases, when the revised reporting rule was implemented; unfortunately, it is not possible to differentiate the numbers of clinically- and laboratory-diagnosed cases.

## Data Availability

The daily cumulative numbers of confirmed and suspected COVID-19 cases for the period from December 1, 2019 to February 8, 2020 were obtained from the National, Hubei Provincial and Wuhan Municipal Health and Family Planning Commissions. The daily population movement data into and out of Wuhan by types of transportation (i.e., long-distance bus, train and air) for the period January 10-23, 2020 were retrieved from an app-derived real-time location database provided by Baidu Huiyan Company.

## Acknowledgements

This work was supported by the National Natural Science Foundation of China from ZJZ(81673239,81973102), WL(11925103 & 61773125), the National Key R&D Program of China (Grant no. 2018YFC0116600), and the STCSM (Grant no. 18DZ1201000).

## Figure legends

**Supplementary Figure 1**: Key parameters regarding COVID-19 epidemic dynamics.

**Supplementary Figure 2**: The decay of COVID-19 controlled reproduction number ***R***_*i*_(***t***) for each province.

**Supplementary Figure 3**: The epidemic peak and final infectious size of each province for COVID-19.

